# Modeling the effects of COVID-19 mobility disruptions on RSV transmission in Seattle, Washington

**DOI:** 10.1101/2024.09.13.24313667

**Authors:** Atchuta Srinivas Duddu, Islam Elgamal, José Camacho-Mateu, Olena Holubowska, Simon A. Rella, Samantha J. Bents, Cécile Viboud, Chelsea L. Hansen, Giulia Pullano, Amanda C. Perofsky

## Abstract

**Introduction:** Respiratory Syncytial Virus (RSV) infection is a major cause of acute respiratory hospitalizations in young children and older adults. In early 2020 most countries implemented non-pharmaceutical interventions (NPIs) to slow the spread of SARS-CoV-2. COVID-19 NPIs disrupted the transmission of RSV on a global scale, and many locations did not experience widespread re-circulation until late 2020 or 2021. Here, we use a mechanistic transmission model informed by cellphone mobility data to determine which aspects of population behavior had the greatest influence on post-pandemic RSV rebound in Seattle, Washington.

**Methods:** We used aggregated mobile device location data to characterize within-city mixing, visitor in-flows, and foot traffic to points of interest in Seattle. We fit an age-structured epidemiological model to data on weekly RSV hospitalizations, allowing for reductions in transmission due to declines in mobility during the pandemic. We compared model fits to observed data to assess which mobility behaviors best capture RSV dynamics during the first two post-pandemic waves in Seattle.

**Results:** In Seattle, COVID-19 NPIs perturbed RSV seasonality from 2020 to 2022. Seattle experienced a small out-of-season outbreak in Summer 2021 and an atypically large and early wave in Fall 2022. RSV transmission models incorporating mobility network connectivity (measured as the average shortest path length between Seattle neighborhoods) or the inflow of visitors from outside of Seattle best captured the timing and magnitude of the first two post-pandemic waves. Models including foot traffic to schools or child daycares or within-neighborhood movement produced poor fits to observed data.

**Conclusions:** Our results suggest that case importations from other regions and local spread between neighborhoods had the greatest influence on the timing of RSV reemergence in Seattle. These findings contribute to the understanding of behavioral factors underlying RSV epidemic spread and can inform the timing of preventative measures, such as the administration of immunoprophylaxis.

## 1 Introduction

In early 2020, many countries implemented non-pharmaceutical interventions (NPIs) to slow the spread of pandemic SARS-CoV-2, including stay-at-home orders, school and business closures, travel bans, and mask mandates. COVID-19 NPIs disrupted the transmission of endemic respiratory pathogens on a global scale, causing low circulation during periods with the most stringent measures, followed by atypically early or out-of-season outbreaks as NPIs relaxed [1, 2, 3, 4, 5, 6, 7, 8].

Many studies have used location data from mobile devices to model the transmission dynamics of SARS-CoV-2 and the effectiveness of control measures ([9, 10, 11]), but relationships between human mobility and the transmission of endemic respiratory pathogens remain less understood. In a previous statistical analysis, increases in population-level mobility preceded or coincided with the post-pandemic rebound of endemic respiratory viruses in Seattle, Washington [8]. However, RSV epidemic dynamics are intrinsically nonlinear, and fluctuations in immunity and susceptibility may obscure relationships between extrinsic factors and epidemic timing [12]. Here, we use a dynamic mathematical model that explicitly accounts for infection and immunological dynamics [12, 13, 14] to explore mechanistic relationships between population behavior and the timing of RSV reemergence in Seattle.

RSV is a major cause of severe lower respiratory tract infections in infants and elderly adults [15, 16] and is responsible for 5% of deaths in children under five [17]. Most children experience their first infection by the age of 2 years, with infants under 6 months at the highest risk for severe infection [18, 19, 20]. RSV protective antibodies wane rapidly, leading to frequent reinfections throughout life, though secondary infections are unlikely to lead to hospitalization [21, 22, 23]. RSV is highly seasonal in temperate regions, with the majority of cases occurring during annual winter epidemics [12]. Climatic and environmental factors are implicated in the seasonality and timing of winter outbreaks [12], while close contacts between young children in daycares and schools drive community transmission [21, 24, 25].

The overall impact of COVID-19 NPIs on RSV circulation is well documented [6, 14, 26, 27, 28], yet the relative contributions of different mobility behaviors to the timing of post-pandemic rebound have not been explored. In this study, we model RSV hospitalizations in a metropolitan county (Seattle-King County, Washington) during pre- and post-pandemic seasons, allowing for fluctuations in transmission due to changes in mobility during the pandemic. To determine specific behaviors that had the greatest influence on the timing of post-pandemic RSV reemergence, we modulate age-specific contacts with different mobility indicators and compare epidemiological model fits to observed RSV dynamics. We specifically focus on mobility metrics that may best approximate transmission-relevant contacts, spatial spread, or external case importations for RSV [8, 25, 29], including foot traffic to schools and child daycares, within-neighborhood movement, the inflow of visitors from outside of Seattle, and the overall connectivity of the Seattle mobility network.

This report describes the results of an educational project conducted within three days’ time during Complexity72h, an interdisciplinary workshop for young researchers in complex systems. Hence, it provides a starting point for future research into the behavioral drivers of RSV epidemics, a topic of great public health importance.

## 2 Methods

Processing of raw mobile device location data, mobility network analyses, and RSV transmission modeling were performed using R version 4.4 and Python version 3.11.

### 2.1 Mobile Device Location Data and Calculation of Mobility Metrics

We obtained mobile device location data from SafeGraph (https://safegraph.com/), a data company that aggregates anonymized location data from 40 million devices, or approximately 10% of the United States population, to measure foot traffic to over 6 million physical places (points of interest) in the United States. We used SafeGraph’s “Weekly Patterns” dataset to estimate the weekly volume of foot traffic to various categories of points of interests (POIs), movement within and between neighborhoods, and the inflow of visitors from outside of King County, WA, during December 2018 - October 2022. POIs are fixed locations, such as businesses or attractions. A “visit” indicates that a device entered the building or spatial perimeter designated as a POI. A “home location” of a device is defined as its common nighttime (18:00-7:00) census block group (CBG) for the past 6 consecutive weeks. We restricted our dataset to King County POIs that had been recorded in SafeGraph’s dataset since January 2019. SafeGraph data were imported and processed using the `SafeGraphR` package [30].

Following [8], we measured movement within and between census tracts (“neighborhoods”) in King County by extracting the home census tract of devices visiting points of interest (POIs) and limiting the dataset to devices with home locations in the census tract of a given POI (within-neighborhood movement) or with home locations in census tracts outside of a given POI’s census tract (between-neighborhood movement). To measure the inflow of visitors from other counties in Washington state or from out-of-state, we limited the dataset to devices visiting POIs in King County with home locations in other WA counties or in other US states, respectively. To measure foot traffic to specific categories of POIs, we aggregated daily visits to POIs by North American Industry Classification System (NAICS) category, without considering the home locations of devices visiting these POIs. To adjust for variation in SafeGraph’s device panel size over time, we divided King County’s census population size by the number of devices in SafeGraph’s panel with home locations in King County each month and multiplied the number of weekly visitors by that value. For each mobility indicator, we summed adjusted weekly visits across POIs to generate county-level mobility indicators.

### 2.2 Weekly Mobility Networks, Degree Centrality, and Shortest Path Lengths

We used the movements of devices residing in King County and visiting POIs outside of their home census tract (i.e., between-neighborhood movement) to create weekly directed mobility networks for the Seattle metropolitan area, wherein individual census tracts (“neighborhoods”) are nodes, and the aggregate movement of devices between census tracts are edges. Edge weights *w*_*ji*_(*T*) were calculated as the sum of the weekly number of trips between each source node *i* and target node *j*.

We used the `NetworkX` Python package [31] to calculate the weighted degree centrality and weighted shortest path lengths among all nodes in the mobility network. We estimated the weighted degree centrality of each node as the sum of edge weights for all edges incident to that node. In each week, we extracted the median weighted degree centrality among all nodes in the network (hereon, *neighborhood degree centrality*). Degree centrality is a measure of node “importance” in the network, wherein nodes with high degree centrality are connected to many other nodes in the network.

To measure path lengths in the network, we considered two nodes to be strongly connected when there is a large flow of devices between them. Given that high edge weights normally indicate large distances between nodes [32], we defined the distance of each link between nodes as the inverse weight along the edge *l*_*ji*_ = 1*/w*_*ji*_. We calculated the shortest path length *L*_*ji*_ between each pair of source node *i* and target node *j* using Dijkstra’s algorithm [32]. Following [33], the mean shortest path length among all nodes *n* in the weekly network is

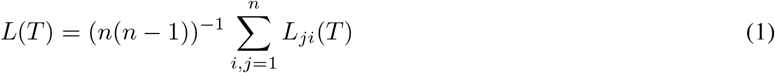

To measure overall *mobility network connectivity* in each week, we inverted mean shortest path lengths *m*(*T*) = 1*/L*(*T*), such that high values correspond to shorter distances (i.e., stronger connections) between nodes.

### 2.3 Hospitalization Data

For individuals ≥10 years, hospital visits diagnosed with RSV in King County, WA were provided by the Rapid Health Information Network (RHINO) program at the Washington Department of Health. For individuals *<*10 years, hospital visits diagnosed with RSV were provided by Seattle Children’s Hospital. We aggregated the two data sources to produce a weekly time series of all-ages hospitalizations spanning January 2017 to April 2023. Due to privacy considerations, weekly hospitalizations with counts between 1-9 were suppressed and re-interpolated. To reduce noise, we applied a centered 3-week moving average to the time series and rounded weekly values to the nearest whole number. Hospitalization rates were estimated as the number of RSV hospitalizations per 100,000 people in King County.

### 2.4 Demographic Data

The initial population size for King County was obtained from 1980 U.S. census data [34]. We assume the birth rate varies over time, according to data on crude annual birth rates from 1980 to 2022. In the transmission model, individuals age exponentially into the next age class, with the rate of aging equal to 1/(width of the age class) [12].

Following Zheng et al. [35] and Hansen et al. [13], the *<*1 year age class is divided into six 2-month age groups. The remaining population is divided into seven age classes: 1 year old, 2-4 years, 5–9 years, 10–19 years, 20–39 years, 40–59 years, and ≥60 years old. The net rate of immigration/emigration and deaths is adjusted to produce a rate of population growth and age structure similar to that observed for King County between 1980 and 2022 [13, 35].

### 2.5 RSV Transmission Model

We use a deterministic age-structured *MSIRS* transmission model for RSV (Maternal Immunity - Susceptible - Infected - Recovered - Susceptible), originally described by Pitzer et al. [12] and Zheng et al. [14], that assumes short-term maternal immunity, frequent reinfection throughout life, and a gradual build-up of partial immunity following infection. Previously, Hansen et al. adapted this model and calibrated it to King County data to project the impacts of new vaccines and extended half-life monoclonal antibodies on RSV hospitalizations during the 2023-2024 season [13]. For the Complexity 72h workshop, we modified Hansen et al.’s model to explore how COVID-19-related behavioral changes influenced RSV transmission dynamics during the first three years of the pandemic, April 2020 - April 2023, prior to the roll-out of new interventions. Model compartments, parameters, and calibration steps are described in detail in a public GitHub repository [13].

Briefly, the model assumes infants are born with transplacentally-acquired antibodies against RSV infection from their mothers (*M*) that wane exponentially within 3-4 months [12, 36]. As maternal immunity wanes, infants become susceptible to infection (*S*_0_). Following each infection (*I*_*i*_), individuals gain partial immunity, which lowers their susceptibility to subsequent infections and the duration and infectiousness of subsequent infections [12, 37, 38, 39]. We assume a progressive build-up of immunity following one, two, three, and four or more previous infections (*I*_*n*_) [21, 22, 23, 40, 12, 14].

The force of infection for a specific age group *a* at time *t*, λ_*a*_(*t*), is influenced by seasonal forcing, age-specific contact rates *c*_*a,k*_, and the total number of infected individuals 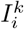 in the population and their relative infectiousness *ρ*:

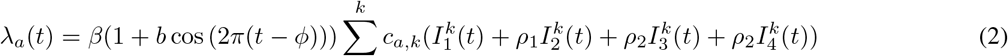

The baseline transmission rate of RSV is *β*, and seasonality in the force of infection is represented by (1 + *b* cos (2*π*(*t* − *ϕ*))), wherein *b* is the amplitude of seasonal forcing and *ϕ* is the phase of seasonal forcing (the timing of peak transmissibility) [12]. The probability of susceptible individuals in age group *a* becoming infected is influenced by their contacts with infectious individuals in the population [12, 13, 29]. *c*_*a,k*_ is the age-specific contact probability between age group *a* and *k* per unit time, and transmission-relevant contacts are assumed to be frequency dependent [12]. The entries of *c*_*a,k*_ were obtained from an expanded version of the age-specific contact matrix described by Mossong et al. [41]. The total number of infected individuals at time *t* is represented by 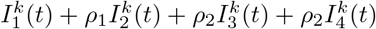 wherein 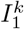 is the number of infected individuals of age *k* experiencing their first infection; 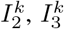, and 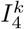 are the number of infected individuals of age *k* experiencing their second, third, or fourth or more infection; and *ρ*_1_ and *ρ*_2_ are the relative infectiousness of the second and subsequent infections compared to the first infection [12, 13, 29]. To link the transmission process to observation-level information, the model estimates a reporting rate that scales infections to reported hospitalizations.

During the pandemic period, we scale the contact matrix *c*_*ak*_ by different time-varying mobility indicators and *µ*_*m*_, a scaling factor estimated by the model. We focus on six mobility metrics that may best approximate transmission-relevant contacts, spatial spread, or external case introductions for RSV: foot traffic to child daycares, foot traffic to elementary and high schools, within-neighborhood movement, the inflow of visitors into Seattle from other Washington counties, mobility network connectivity, and neighborhood degree centrality. We also tested the inflow of out-of-state visitors but this metric did not improve model fit (see *Discussion*). Prior to inclusion in the model, mobility metrics were re-scaled to fall between 0 and 1, such that values close to 0 occur when mobility is low (e.g., the stay-at-home period), and values close to 1 occur when mobility is at “typical” pre-pandemic levels. We assume that contact rates are at baseline levels from January 2017 to March 2020, are modulated by changes in mobility during the pandemic, from April 2020 to March 2022 (the start of stay-at-home measures to the end of the Omicron BA.1 wave), and linearly return to baseline by Fall 2022.

We use maximum likelihood estimation (MLE) to fit the model to the pre-pandemic and pandemic periods (see [13] for details). For each time period, the likelihood of the data given the model is estimated by assuming the number of hospitalizations in each age class *a* during each week is Poisson-distributed with a mean equal to the model-predicted number of RSV infections, multiplied by the reporting rate [13, 12, 14]. We use the `deSolve` R package [42] to solve the *MSIRS* model. To perform model selection, we use Akaike Information Criterion (AIC) [43] to measure model fits to observed hospitalizations.

### 2.6 Ethics Oversight

The study was determined to be non-human subjects research by the University of Washington Institutional Review Board (#00017840). The human cellphone mobility data are aggregated and anonymous and were freely available to academic researchers prior to the start of this study; thus, these data do not constitute human subjects research. There is no individual-level linkage between the hospitalization and mobility datasets. Individual-level linkage between these two datasets is not possible, given that both datasets are aggregated and anonymous/de-identified.

## 3 Results

### 3.1 Disruptions to population mobility during the COVID-19 pandemic

The first community acquired COVID-19 case in the United States was reported in the Seattle metropolitan area on February 28, 2020. To slow the spread of SARS-CoV-2, Washington state declared a State of Emergency of February 29, 2020, closed schools in King County on March 12, and enacted statewide stay-at-home orders on March 23. King County also recommended that workplaces allow employees to work from home on March 4 and closed indoor dining and other businesses on March 16.

We used aggregated mobile device location data to approximate contact and movement patterns that are potentially relevant for RSV transmission (Figure 1). Population mobility in King County declined dramatically after the State of Emergency in late February 2020 (Figure 1). Visitor inflow, mobility network connectivity, and median neighborhood degree centrality began to increase after the lifting of stay-at-home orders in early June 2020, gradually rose throughout 2020, and reached pre-pandemic levels in early 2021. Foot traffic to schools rose slightly after Washington required public schools to provide in-person instruction in April 2021 and fully returned to pre-pandemic levels during Fall 2021 (Figure 1). Within-neighborhood movement and foot traffic to child day cares were still below pre-pandemic levels by the end of our mobility dataset in October 2022 (Figure 1).

**Figure 1:**
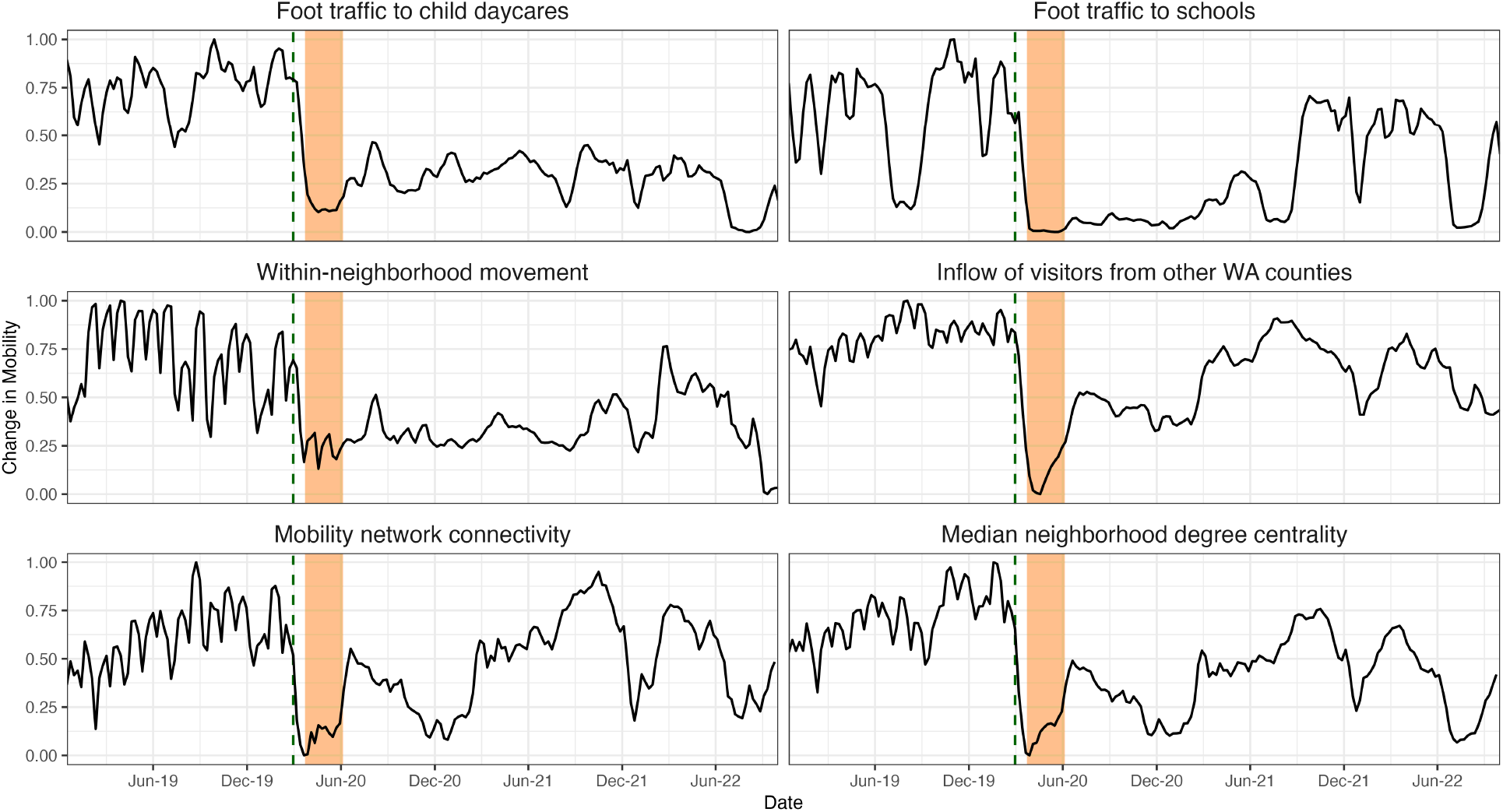
Weekly mobility levels in Seattle-King County, Washington based on aggregated mobile device location data from SafeGraph, December 2018 – October 2022. In each panel, the vertical dashed line indicates the date of Washington’s State of Emergency declaration (February 29, 2020), and the vertical orange shaded panel indicates Seattle’s stay-at-home period (March 23 – June 5, 2020).

### 3.2 RSV hospitalization patterns during the COVID-19 pandemic

In the greater Seattle region, the transmission rates of all respiratory pathogens dropped substantially after the State of Emergency on February 29, although the winter RSV epidemic had mostly subsided by late February 2020 [8]. COVID-19 NPIs disrupted RSV transmission throughout 2020 and 2021, and RSV did not widely re-circulate in Seattle until Summer 2021. Seattle experienced a small, out-of-season wave of RSV hospitalizations beginning in June 2021 (Figure 2), coinciding with increasing visitor inflow and between-neighborhood movement, declines in masking, and the return to in-person learning for school students [8]. RSV hospitalizations continued to rise throughout Fall 2021, peaked in mid-December 2021, and declined rapidly during the Omicron BA.1 wave in January 2022 (Figure 2). During Fall 2022, Seattle experienced a second, atypically large wave of RSV hospitalizations that peaked in early November, two months earlier than the seasonal average for pre-pandemic RSV seasons (Figure 2).

**Figure 2:**
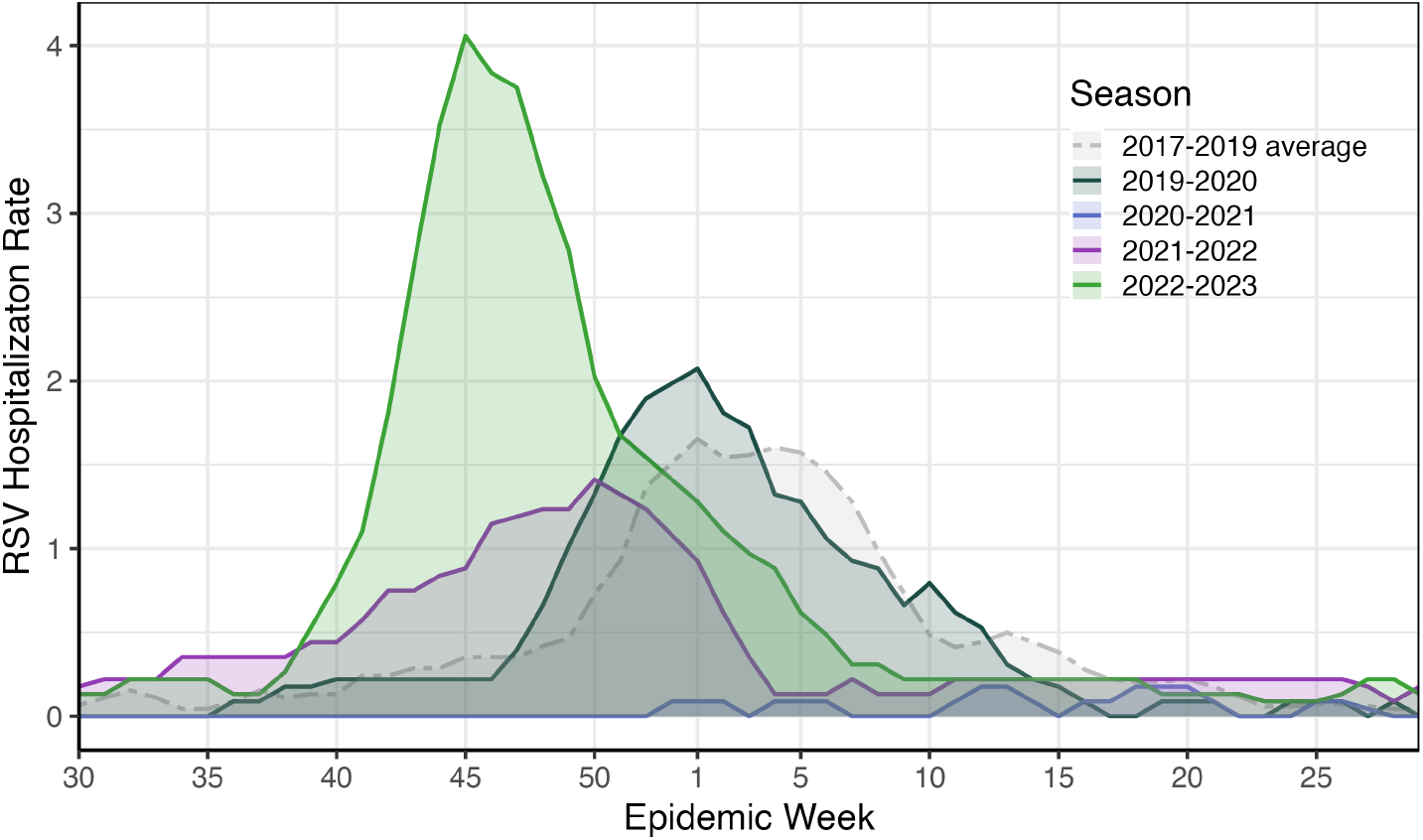
Seasonal RSV hospitalization rates in Seattle-King County, Washington, during January 2017 - July 2023. Weekly hospitalization rates during the 2019-2020, 2020-2021, 2021-2022, and 2022-2023 respiratory virus seasons are compared to the average weekly rate of three pre-pandemic seasons (2017-2019). Hospitalization rates were estimated as the number of RSV hospitalizations per 100,000 people in Seattle.

### 3.3 Modeling the impact of population mobility on RSV hospitalizations during the COVID-19 pandemic period, 2020 - 2023

During the pandemic period, we observed considerable variation across epidemiological model fits to observed RSV hospitalizations, depending on the particular mobility metric modulating contacts in the model. The model incorporating mobility network connectivity best captured the timing and magnitude of the two waves of RSV hospitalizations in Summer 2021 and Fall 2022, respectively, though it overestimated the size of the first peak and slightly underestimated the size of the second peak (Figure 3). The inflow of visitors from other counties in Washington reproduced the general timing of both waves and the size of the first wave in 2021, but underestimated the magnitude of the second wave in 2022. Foot traffic to schools and neighborhood degree centrality also reproduced the timing and size of the first wave in 2021 but inaccurately estimated a gradual increase in hospitalizations throughout 2022, instead of a distinct second wave during Fall 2022 (Figure 3). Lastly, models including within-neighborhood movement or foot traffic to child daycares entirely missed the first wave and substantially overestimated the second wave (Figure 3).

**Figure 3:**
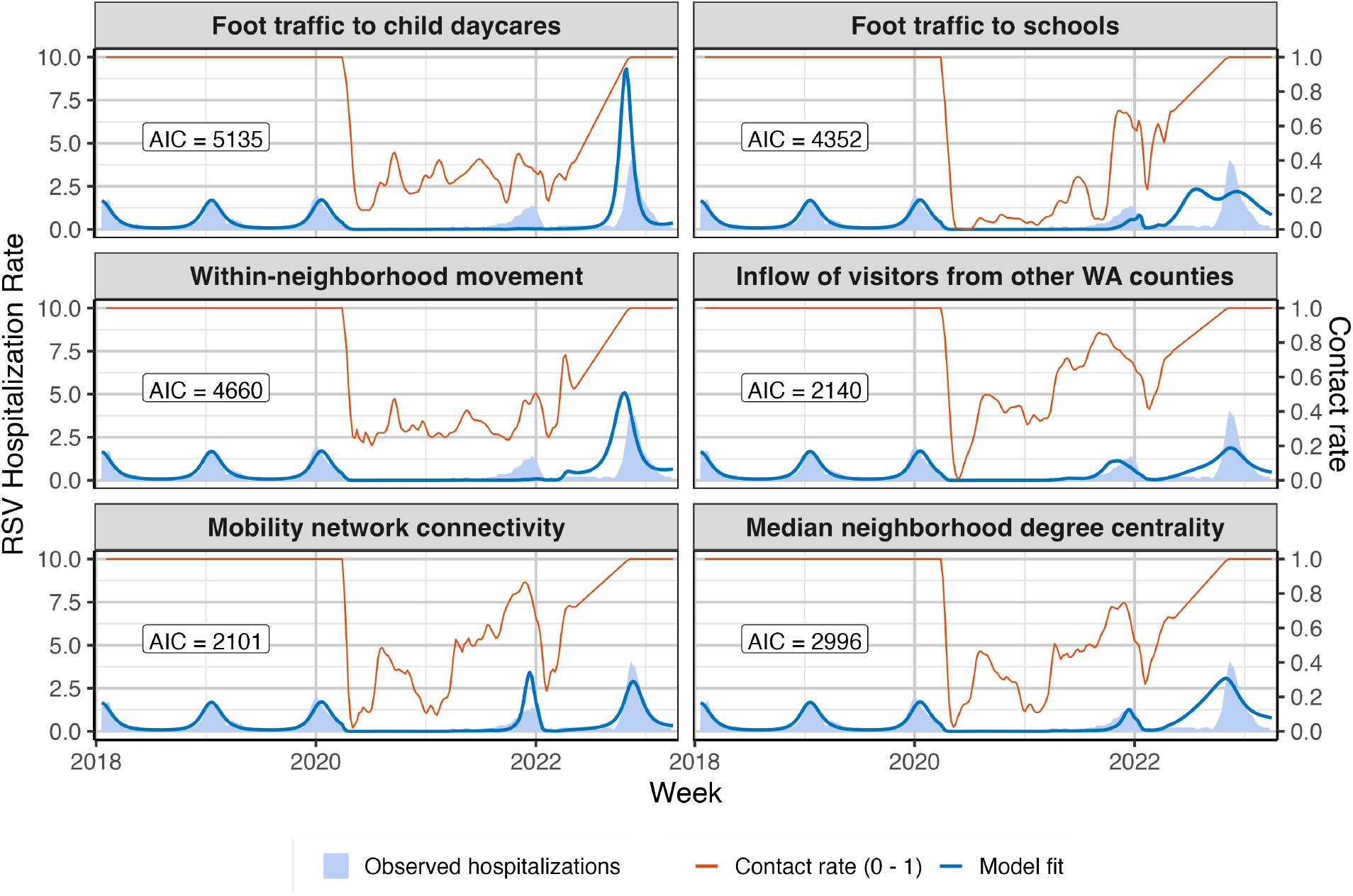
Transmission model fits to reported weekly RSV hospitalization rates in Seattle-King County, Washington, January 2017 - April 2023. Age-structured models of RSV transmission were fit to observed hospitalizations, allowing for reductions in contacts due to physical distancing during the COVID-19 pandemic. In separate models, we scaled the age-specific contact matrix by six time-varying mobility metrics: foot traffic to child daycares, foot traffic to elementary and high schools, within-neighborhood movement, the inflow of visitors from other counties in Washington state, mobility network connectivity, and median neighborhood degree centrality. Contact rates were assumed to be at baseline levels from January 2017 to March 2020, were adjusted by mobility changes from April 2020 to March 2022 (the end of the Omicron BA.1 wave), and linearly returned to baseline during Fall 2022. We used AIC to measure goodness of fit of models to observed data, with smaller AIC values indicating better model performance.

Among candidate models, the model incorporating mobility network connectivity best fit observed hospitalizations (lowest AIC score), followed by the model informed by the inflow of visitors from other Washington counties (Table 1). Models including the remaining mobility indicators had comparatively poorer fits to the data (Table 1).

**Table 1:** Likelihood-based comparison of RSV transmission models incorporating mobility.

| Model | Log-likelihood | No. of parameters | AIC | ΔAIC |
| --- | --- | --- | --- | --- |
| Mobility network connectivity | -1048 | 2 | 2101 | 0 |
| Inflow of visitors from other WA counties | -1068 | 2 | 2140 | 39 |
| Neighborhood degree centrality | -1496 | 2 | 2996 | 895 |
| Foot traffic to schools | -2174 | 2 | 4352 | 2251 |
| Within-neighborhood movement | -2328 | 2 | 4660 | 2559 |
| Foot traffic to child daycares | -2566 | 2 | 5135 | 3034 |

## 4 Discussion

COVID-19 mitigation measures had profound effects on the transmission dynamics of endemic infectious diseases, including respiratory syncytial virus (RSV). Here, we calibrate a mechanistic, compartmental model of RSV transmission to multiple years of hospitalization data in Seattle-King County, Washington and model the impacts of COVID-19 NPIs and associated behavioral changes on post-pandemic RSV reemergence.

### 4.1 RSV dynamics during the first years of the COVID-19 pandemic

Hospitalization data and modelling results show that COVID-19 NPIs significantly perturbed RSV seasonality from 2020 to 2022. The first year of the pandemic was characterized by strict NPIs, high levels of SARS-CoV-2 circulation, and very low to no RSV circulation. Although Seattle stay-at-home orders lifted in early June 2020, RSV did not widely re-circulate until the summer months of 2021, likely due to Seattle residents’ strong adherence to social distancing measures and high rates of mask wearing throughout 2020 and 2021 [8].

Beginning in June 2021, Seattle experienced a small “off-season” wave of RSV hospitalizations that peaked in December 2021 and sharply declined in January 2022, coinciding with a surge of COVID-19 cases associated with the novel Omicron BA.1 variant. Low population exposure to RSV during 2020 and 2021 is expected to have led to an accumulation of infants susceptible to primary infection and waning of immunity in previously exposed individuals [26]. Despite this buildup of susceptible individuals, the first post-pandemic wave of RSV in Seattle was small compared to pre-pandemic seasons, likely due to many COVID NPIs still being in place during 2021 [6, 14, 44]. The precipitous drop in RSV hospitalizations during January 2022 may have resulted from Seattle residents staying home to avoid exposure to the highly transmissible Omicron variant or due to negative interference between the Omicron virus and endemic respiratory viruses [8].

During Fall 2022, Seattle experienced a much larger, more severe wave of RSV hospitalizations that began in September and peaked in early November, two months earlier that the average timing of pre-pandemic RSV seasons. Because the 2021 wave was smaller than those in typical RSV seasons, many infants and young children were likely still unexposed to RSV going into the 2022-2023 season. This “immunity debt”, compounded by relaxed NPIs, may explain the early timing and increased severity of the 2022 epidemic. Although we do not examine the age distribution of RSV hospitalizations in this report, several studies have documented a shift in RSV hospitalizations to older infants after the relaxation of COVID-19 NPIs [6, 13, 45, 46].

### 4.2 The role of human mobility in post-pandemic RSV reemergence

To determine which aspects of population behavior had the greatest influence on RSV reemergence, we fit epidemiological models to observed hospitalizations, allowing for fluctuations in transmission due to changes in population mobility during the pandemic. Models incorporating mobility network connectivity (the average shortest path length between neighborhoods in Seattle), followed by the inflow of visitors into Seattle, best reproduced the timing and magnitude of the first two post-pandemic RSV waves. These preliminary findings suggest that seeding from external sources and viral dispersal between neighborhoods had the greatest impact on RSV rebound in Seattle.

First, cellphone data show that the inflow of visitors from outside of Seattle was 50% below baseline throughout 2020 and did not return to pre-pandemic levels until late spring or summer 2021 [8]. It is well established that annual influenza epidemics in North America are seeded via air travel by strains originating in East and Southeast Asia [47], but the contribution of local persistence versus external seeding to RSV epidemics is not well understood [12, 14]. Our modelling results suggest that increasing visitor inflow in 2021 imported RSV cases from other regions, seeding new outbreaks in Seattle [48, 14, 8]. In particular, visitor inflow from other counties in Washington fit observed epidemic dynamics better than visitor inflow from other states (not shown). We speculate that regional travel may better approximate the local movements of children than the influx of out-of-state visitors, a proxy for air travel [47]. Additionally, the peak timing of out-of-state visitor inflow during summer months does not coincide with the winter seasonality of RSV epidemics.

Second, COVID-19 NPIs led to reduced travel between neighborhoods [8], fracturing the Seattle mobility network and in turn slowing the spatial spread of infectious diseases [33]. Our findings indicate that the reformation of links between Seattle neighborhoods may have been key in facilitating within-city viral dispersal after RSV was reintroduced into Seattle.

Surprisingly, models incorporating proxies for the movements of children - foot traffic to schools or child daycares - produced overall poor fits to observed hospitalizations. For example, the model including foot traffic to schools generally reproduced the timing and size of the first post-pandemic wave but vastly overestimated the second wave of hospitalizations. Although children are the primary transmission group for RSV, and school reopenings are linked to increased RSV activity, our study and others suggest that school reopenings cannot fully explain the timing of post-pandemic RSV reemergence [49, 50]. Our results are also consistent with studies of pre-pandemic seasons that found school-term forcing does not explain RSV epidemic timing [12, 29, 51]. Further research is needed to determine mobility metrics that may be better proxies for school-based contacts and other points of interest relevant to RSV transmission.

### 4.3 Limitations

Our study is subject to several limitations. First, prior modelling studies show that accounting for interactions between RSV subtypes explains epidemic dynamics in the United Kingdom and Finland [37, 44]. However, our model does not consider the effects of RSV-A and RSV-B subtype interference on transmission. In Seattle, the first post-pandemic wave was comprised of RSV-B cases, whereas the second wave was dominated by RSV-A [8]. Prior to the pandemic, both subtypes co-circulated during winter epidemics [8]. Although we speculate that the more severe second wave of RSV hospitalizations resulted from the combined effects of “immunity debt” and decreased social distancing, RSV-A’s higher transmissibility and tendency to cause more severe infections in young children may have also contributed the second wave’s larger size [44]. Second, young children are more susceptible to RSV infection than older age groups, and close contacts among children are considered to be the primary driver of RSV transmission in the community. However, SafeGraph does not track individuals younger than 16 years of age. Our inability to directly measure the movements of children could obscure the relationship between school-based contacts and RSV transmission. Third, relationships between cellphone mobility and infectious disease transmission are stronger in urban cities than in rural locations [52, 53, 54]. Because our study is limited to a single metropolitan area, our findings may not be applicable to more sparsely populated counties. Lastly, we did not fit models to the 2023-2024 RSV season because the roll-out of new RSV vaccines and extended half-life monoclonal antibodies introduces complexities beyond the scope of this study.

## 5 Conclusions

Although our study is preliminary, it suggests that human mobility patterns are an important driver of RSV spread. Our initial findings indicate that the importation of external infections and spatial spread between neighborhoods influenced the timing of post-pandemic RSV reemergence in Seattle. Understanding the effects of mobility behavior on RSV epidemics is important for clinical and public health preparedness. Insights from this research could ultimately inform the timing of preventative measures for RSV, such as the administration of long-acting monoclonal antibodies or vaccines. Ongoing work is focused on refining the inclusion of mobility data in transmission models and calibrating the model to observed age distributions of hospitalizations.

## Data Availability

Raw hospitalization data for King County, WA are not publicly available but the final dataset of hospitalizations used in this study can be accessed at https://github.com/chelsea-hansen/RSV-Interventions. Aggregated mobility data for King County can be accessed at https://github.com/aperofsky/seattle_mobility_rt. The SafeGraph Weekly Patterns dataset is available to academics for non-commercial use through an institutional university subscription or individual subscription to Dewey (https://www.deweydata.io/). The data access agreement with Dewey does not permit sharing of the raw data.

https://github.com/aperofsky/seattle_mobility_rt

## 6 Additional Information

## Acknowledgements

This work is the output of the Complexity72h workshop, held at the Universidad Carlos III de Madrid in Leganés, Spain, 24-28 June 2024 (https://www.complexity72h.com). A.C.P. acknowledges support from the Fogarty International Center at the U.S. National Institutes of Health to carry out this project.

This study modifies a transmission model for RSV originally developed by C.L.H. in collaboration with Public Health - Seattle & King County [13] for an initiative supported by the U.S. Council of State and Territorial Epidemiologists and U.S. Centers for Disease Control and Prevention: “Development of forecast, analytic, and visualization tools to improve outbreak response and support public health decision making.”

We thank the Washington Department of Health, Public Health - Seattle & King County, and Seattle Children’s Hospital for providing RSV hospitalization data, and Lawrence Lee for facilitating access to these data. We thank SafeGraph and Dewey for providing access to mobility data. We also thank Daniela Paolotti for helpful discussions about the design of the project.

## Disclaimer

The findings and conclusions in this report are those of the authors and do not necessarily represent the official position of the U.S. National Institutes of Health or the U.S. government.

## Competing Interests

C.L.H reports receiving personal fees from Sanofi outside the submitted work. C.V. reports receiving honoraria from Elsevier outside the submitted work. No other disclosures were reported.

## Data Availability

Raw hospitalization data for King County, WA are not publicly available but the final dataset of hospitalizations used in this study can be accessed at https://github.com/chelsea-hansen/RSV-Interventions [13]. Aggregated mobility data for King County can be accessed at https://github.com/aperofsky/seattle_mobility_rt [8]. The SafeGraph Weekly Patterns dataset is available to academics for non-commercial use through an institutional university subscription or individual subscription to Dewey (https://www.deweydata.io/). The data access agreement with Dewey does not permit sharing of the raw data.

